# Modelling the urban acoustic environment using land use-based gradient boosting

**DOI:** 10.1101/2025.07.22.25331920

**Authors:** Timo Haselhoff, Susanne Moebus, Mikel Jedrusiak, Bryce T. Lawrence, Frank Weichert

## Abstract

**Background:** Long-standing research on the relationship between the urban acoustic environment (AE) and human health demonstrates the harmful effects of environmental noise. Meanwhile, an increasing number of smaller studies report health benefits for additional acoustic properties. However, studies on health-promoting AEs remain limited, largely due to the lack of methods for estimating high-resolution acoustic properties beyond conventional noise metrics.

**Objective:** We investigate to what extent models based on land-use types (LUT) can predict properties of the urban AE, focusing on four acoustic indices (Articulation Index, Bioacoustic index, Link Density and Sharpness). Additionally, we predict the LAeq, which enables us to compare the performance between our model, the strategic noise map of Bochum (SNM) and results from the literature.

**Methods:** We use a dataset of 2,746 acoustic measurements from 785 locations in Bochum and 22 locations in Essen (n=90) to train and evaluate gradient boosting models. For model development, data is split into training/validation-(668 locations in Bochum) and test-sets (117 locations in Bochum and all locations in Essen). The models predict acoustic indices based on the area of 77 LUTs within 50 and 300 m buffers around each location.

**Results:** Based on the root mean square error (RMSE), predictions for Link Density deviate on average by 0.17 and 0.21 from test-sets in Bochum and Essen. For the LAeq, the RMSE is 4.8 dB(A) and 4.4 dB(A), respectively. The R^2^ for the Link Density is between 0.27 and 0.3, and for the LAeq between 0.52 and 0.46. The SNM performs worse in predicting the LAeq (RMSE=7.8; R^2^=-0.31). Performances for other indices are mixed.

**Significance:** LUT-based models demonstrate their potential for predicting Link Density and LAeq, achieving moderate to strong performance across two independent test datasets. This provides a scalable approach for investigating potentially health-relevant properties of the urban AE at high spatial resolution.

## 1. Introduction

Considerable research efforts have been made to investigate the relationship between the urban acoustic environment (AE) and human health. The adverse health effects of environmental noise exposure (e.g. cardiovascular disease and mental health issues) are well documented [1]. However, the AE can be defined as “the sound from all sound sources as modified by the environment” [2], encompassing a broader range of acoustic properties than environmental noise alone (e.g. underlying sound sources, temporal variations or frequency characteristics). A growing body of research is now examining the potential health benefits of additional acoustic properties [3]. Laboratory studies, for instance, have shown that calm and pleasant sounds are associated with greater reductions in heart rate [4], that natural sounds can promote stress recovery [5], and that biophonic AEs can positively influence functional brain connectivity [6]. Beyond laboratory settings, several field studies have also investigated the impact of natural sounds on various aspects of human health, reporting associations with reduced pain, lower stress levels, and improved cognitive performance [7].

However, there is a significant lack of comprehensive population-based studies examining the relationship between the AE and human health beyond environmental noise, which are crucial for examining the patterns, causes, and effects of health outcomes in defined population groups within real-life settings [8] (pp. 555-583). Such studies require high spatial resolution data to assess exposure to the respective acoustic properties under investigation. For research on the relation between environmental noise exposure and human health, data is already available through strategic noise maps (SNMs), mandated by the Environmental Noise Directive (END) in the EU [9]. Here, SNMs with high spatial resolution are created for agglomerations of over 100,000 inhabitants, as well as for major roads, railways, and airports. The SNMs assess exposure to “unwanted or harmful outdoor sound created by human activities” [9]. While they are effective at quantifying noise emitted by “transport, road traffic, rail traffic, air traffic, and from sites of industrial activity” [9], there is currently no established method to estimate exposure to additional acoustic properties of the AE at a comparable spatial resolution. Consequently, the current unavailability of high spatial resolution data on the AE beyond environmental noise exposure is a major barrier to conducting population-based studies on the health impacts of the wider AE.

A promising approach to overcome this barrier of exposure assessment involves the use of land use-based models. These models can estimate environmental exposures at high spatial resolution by leveraging the statistical relationships between measured exposure data (e.g. air pollution) and land use types (LUTs) (e.g. highways or green space). Land use-based models were already successfully used for the estimation of outdoor air pollution [10]. The applicability of such models to acoustics is demonstrated by their effective use in estimating average noise levels, traffic noise, and in the extrapolation of SNMs [11-19]. Some of the major advantages in the application of LUT based models is that they are cost-effective and computationally inexpensive, especially when compared to more complex alternatives like the creation of SNMs [17]. In addition, they are easily scalable as LUT information is often readily available and thus suited for data-poor regions where detailed exposure measurements are limited or unavailable (e.g. agglomerations with less than 100,000 inhabitants). Furthermore, as they are based on land use information, they can be directly integrated into urban planning processes. However, to the best of the knowledge of the authors, land use-based models have not yet been applied to estimate AE properties beyond noise.

In this work, we investigate the potential of LUT based models to estimate acoustic properties beyond environmental noise exposure, as defined by the END. We focus on the acoustic properties captured by the Articulation Index [20], the Bioacoustic Index [21], the Link Density [22] and the maximum Sharpness [23]. These indices were chosen, as previous works have already shown their relations to LUTs as well as the human perception of pleasantness [24], health related urban greenspace [25] and biophonic activity [21, 26], thus, indicating potentially health relevant acoustic properties. In addition, we integrate the A-weighted sound pressure level (LAeq) as a measure of total environmental noise. This also enables us to compare the performance to predict total environmental noise between our model, the SNM as well as results from the literature. To facilitate a scalable approach, we rely exclusively on LUT as predictor variables because they are usually easily accessible and, in many cases, even legally mandated. Specifically, the research questions are:

1. To what extent can gradient boosting models based on LUTs predict properties of the urban AE at high spatial resolution?
2. How well do these models perform in estimating:
  i. Acoustic properties beyond noise, when tested on same-source data and external-source measurements?
  ii. Total environmental noise levels, when tested on same-source data, external-source measurements and in comparison to SNM predictions?

## 2. Material and Methods

To address our research questions, we train a gradient boosting model on LUT data provided by local authorities and acoustic measurements from the SALVE (AcouStic QuAlity and HeaLth in Urban EnVironmEnts) project in Bochum [27]. We evaluate the performance of the model using 5-fold cross validation, test data from SALVE as well as acoustic measurements from the Be-MoVe (Participation-based transformation of active mobility for health-promoting urban and transport infrastructures) project conducted in Essen, Germany [28]. Additionally, we compare the model’s performance in estimating total noise levels with the performance of the SNM of Bochum. Finally, we demonstrate the application of the model to the area of Bochum, Essen and the neighbouring city of Mülheim an der Ruhr.

### 2.1 Data on the acoustic environment

#### SALVE

To train and test our model, we rely on acoustic measurements from the SALVE project in Bochum. A comprehensive description of the SALVE study design is provided in Haselhoff et al. [27]. The sampling strategy was designed to capture the environmental contexts in close proximity to people’s homes. The recording location selection was based on stratified LUT sampling as described in [27]. At each sampling point, four audio recordings (5-min, 48 kHz, 24-bit depth) were made once each season between 03.2019 and 03.2020. Recordings took place between 09:00 and 17:00. The recording device was a NTi XL2 sound recorder with the M2230 omni-directional microphone [29], mounted at a height of around 1.65 m. The device was calibrated to meet the standards IEC 61672:2013, IEC 61672:2003, IEC 61260:2014, IEC 61260:2003, IEC 60651 and IEC 60804. A total of 2,746 audio recordings were gathered at 785 locations. The number of recording locations differs from 730, as we considered measurement location deviations of more than 5 m as significant.

#### Be-MoVe

To test our model, we draw data from the Be-MoVe project, which aimed to test co-created alternative forms of mobility and designs of public spaces in urban neighbourhoods. The study design is described in detail in Hornberg et al. [28]. Shortly, as part of the Be-MoVe project, soundwalks according to DIN ISO 12913 [2] were conducted to assess the acoustic environment at 22 locations in two districts in Essen. The districts are characterised by (i) apartment buildings, recreational areas and open space and (ii) diverse urban landscape, ranging from shopping streets and commercial centres to densely populated residential zones. For each soundwalk, at each listening station, an acoustic measurement (3-min, 48 kHz, 24-bit depth) was conducted, using the same device as within the SALVE project. Recordings were made between 10.05.2023 and 06.10.2023 between 18:00 and 19:30. In total, we completed 90 measurements across 22 listening stations.

#### Acoustic Properties

For each recording, a number of acoustic indices are derived: The total noise level is assessed using the A-weighted sound pressure level (LAeq), which was directly provided by the recording devices. As a measure of intelligibility of human voice, we use the Articulation Index based on the root mean square (RMS) of the signal. The Articulation Index indicates how much background noise level can interfere with human speech and ranges from 0 (no speech understood) to 1 (all speech understood) [20]. As a measure of the energetic centre of gravity between frequencies, we use the maximum Sharpness value during each recording, according to the DIN 45692 [23]. It is measured in Acum and ranges from zero to infinity, where higher values indicate sounds with more energy in higher frequency bands (i.e. whether a sound is perceived as sharp, shrill, bright or hissing). The Articulation Index as well as the Sharpness are calculated using ArtemiS SUITE (version 15.1). Additionally, we use the Bioacoustic Index (BIO). The BIO ranges from zero to infinity, where higher values indicate greater differences between the quietest and the loudest 1 kHz frequency band in-between 2 and 8 kHz. In more rural areas, higher values shall indicate higher avian abundance, while in urban areas, higher values are found to be more indicative for road traffic and less green space [21, 26]. The BIO is calculated following the script used in Lawrence et al. [30]. Furthermore, we include the Link Density as measure of acoustic dominance, i.e., a measure of how many factors (i.e. sound sources) contribute to the overall spectral dynamic. It ranges from zero to one, where higher values indicate higher acoustic dominance (e.g. by cars passing by). The Link Density is calculated following the procedure outlined in Haselhoff et al. [25].

As we want to predict the average acoustic properties at each location, we calculate the mean of all indices (using energetic averages for the LAeq) for each location from the measurements conducted at each sampling point, resulting in 785 measurement locations for the SALVE and 22 for Be-MoVe.

#### Training, Validation and Test Split

We partition the SALVE dataset into a training/validation (85%, n = 668) and a test (15%, n = 117) subset, orientating us at Almansi et al. [31] and the commonly used 70/30 split between training/validation and test data [32]. We call the latter test split SALVE_Test_. The training/validation data are used for model development through 5-fold cross validation. The SALVE_Test_ subset and the Be-MoVe dataset are used to evaluate the models performance on unseen data.

#### Strategic Noise Maps

To evaluate our model’s performance in predicting total environmental noise, we additionally compare it to the performance of the SNM in Bochum. For this, we received the SNM from the city of Bochum from 2022. To be comparable to our recordings, we use the L_Day_ in 1 dB(A) increments, i.e., the A-weighted long-term average sound level between 06:00 and 18:00, to match our recording times. The provided values range from 34.5 to 79.5 dB(A). Since there are multiple SNM depicting different noise sources (road traffic, railway traffic and industry), we combine them by overlaying the maps and energetically summing their respective LAeq values. We compare the LAeq measurements from SALVE with corresponding values from the SNM by extracting the predicted LAeq from the SNM at each measurement location.

### 2.2 Environmental Data

The predictor variables are based on land use data provided by the Ruhr Regional Association (RVR) for the year 2019 [33]. In total, there are 146 LUT categories. We derive the LUTs around each recording location by calculating the proportion of each LUT category within buffers of (i) 50 m, to capture the immediate surrounding of each location, and (ii) 300 m radius, to capture potentially important acoustic impacts of LUT within a wider distance. Altogether, this results in 292 possible predictor variables per recording location.

## 2.3. Methods

### Descriptive Statistics

To provide an overview on the acoustic properties of our recordings, we show descriptive statistics, including the arithmetic mean, the minimum, the maximum and the standard deviation of the SALVE and the Be-MoVe datasets. In addition, since we have multiple recordings for each location, we report the within-location standard deviation to capture the inherent variability of respective acoustic properties. The reported within-location standard deviation is the arithmetic mean of each within-location standard deviation per recording location.

### Feature Reduction

To improve the efficiency of the gradient boosting models, we remove non-related LUT categories from the set of predictor variables by using Boruta feature selection. Briefly, Boruta feature selection is a method that identifies all relevant features by comparing the importance of the actual variables to that of randomly permuted “shadow features” of these variables, using a random forest classifier [34]. We repeat this for each acoustic index against all 292 LUT variables in our dataset and keep those LUT variables, which were identified by the algorithm as “important” or “tentative”. Finally, we harmonize all identified LUT variables into a single predictor set for all acoustic indices (Appendix). We perform the calculation using the “Boruta” function from the Boruta package (version 8.0.0) in R (4.1.3), applying the default settings as recommended by the authors [34].

### Gradient Boosting

We train five separate gradient boosting regressor models to predict the respective acoustic indices using the LUT features identified by the Boruta algorithm [34]. Gradient boosting builds a predictive model by sequentially combining multiple weak learners. The gradient boosting framework consists of three key components: (i) a loss function to be minimized, (ii) a weak learner for generating predictions, and (iii) an additive model that incorporates new learners to reduce the residual errors of the ensemble. There are three main hyperparameters, which impact the performance of the model: the number of trees, the maximum depth of the trees, and the learning rate [35]. We optimize these parameters by performing a grid search combined with a 5-fold cross validation. A 5-fold cross-validation is an evaluation technique used to assess how well a model generalizes to unseen data. The dataset is randomly divided into five equal parts (folds). In each of five iterations, four folds are used to train the model, and the remaining fold is used for validation. This process repeats until each fold has been used once for validation. The final performance metric is the average of the five validation results. For the grid search, we iterate over the hyperparameters number of trees (10, 50, 100, 500, 1000, 5000), maximum tree depth (3, 4, 5, 6), and learning rate (0.0001, 0.001, 0.01, 0.1, 1.0). For each hyperparameter combination, a 5-fold cross-validation is used to evaluate the model performance. The optimal hyperparameter combination is selected based on the lowest root mean square error (RMSE). In the following, we call the model based on 668 locations SALVE_Train_. Once the optimal hyperparameters are defined, and after the model is tested against the SALVE_Test_ data, we train the model on the entire SALVE dataset to maximize its learning from all available data and prepare it for the evaluation on the Be-MoVe data. This model is referred to as SALVE_All_.

### Performance Measures

To evaluate the performance of each model, we mainly rely on three performance measures: The root mean square error (RMSE), the mean absolute error (MAE) and the coefficient of determination (R^2^). RMSE provides a measure of the average magnitude of the prediction error, with greater weight given to larger errors. MAE quantifies the average absolute difference between predicted and observed values, offering a more interpretable and less sensitive measure to outliers compared to the RMSE. R^2^ indicates the proportion of variance in the observed data explained by the model, with values closer to 1 signifying better predictive performance. In addition, we provide a visualisation to compare the model predictions against the true values using scatter plots as well as histograms.

### Shap

For a more detailed investigation of the model’s performance, we assess the importance of each predictor variable using SHAP (SHapley Additive exPlanations). SHAP values provide a unified measure of feature importance by quantifying the contribution of each variable to individual predictions [36]. This approach allows for a consistent and interpretable explanation of the model output by attributing the prediction difference from the mean to each feature. We report SHAP values using a beeswarm plot for the five most important predictors for each model. SHAP values are derived by the application of the SALVE_All_ model on the entire SALVE dataset.

### 2.4 Sound Maps

To demonstrate the scalable properties of the LUT-based model, we generate “sound maps” for the cities of Bochum, Essen, and Mülheim. If the models demonstrate sufficient performance, they could enable population-level exposure estimates to AE properties beyond conventional environmental noise metrics. To construct the sound maps, we overlay a grid of equally spaced sampling points (30 m apart) across the three cities. For each point, we calculate the LUT variables used in the model within buffers of 50 m and 300 m. We then apply the final prediction model to estimate acoustic properties at each location. To enhance the visualization, we use Kriging [37] to interpolate values between the sampling points, creating a continuous surface of predicted noise levels.

We calculate gradient boosting, hyperparameter tuning and performance measures using the sklearn package (0.42.2) and SHAP values using the shap package (0.46.0) in python (3.9.7). The calculation of LUT areas in two buffer sizes, the sampling of the sound map points and the Kriging are performed using ArcGIS (3.2.0).

## 3. Results

### 3.1 Data Description

The LAeq for the SALVE dataset has a mean of 55.306 dB(A) with a range between 33.584 and 79.260 dB(A) (Table 1). The standard deviation (STD) is higher between recording locations than within recording locations, showing a more stable sound pressure level at each location than between locations. In the Be-MoVe dataset, the STD is also higher between recording locations (5.066 dB(A)) than within them (2.501 dB(A)), but the values are overall lower here, underlining a less diverse AE in terms of LAeq than the one measured in the SALVE project. The Articulation Index has a mean of 0.846, a minimum of 0.207 and a maximum of 1 in the SALVE dataset, while also having a much higher between (0.136) than within STD (0.076). This tendency is even stronger in the Be-MoVe dataset, with a between STD of 0.146 and a within STD of 0.051. In contrast, the between STD for the BIO is smaller than the within STD in both datasets (SALVE: 0.985, 1.190; Be-MoVe: 0.824, 1.249), showing a higher variation within recording locations than between. For the Link Density, this behaviour is also observable for the SALVE dataset, but much less pronounced with a between STD of 0.203 and a within STD of 0.210. Furthermore, the mean is higher in SALVE with 0.563 against 0.478 in Be-MoVe and the Be-MoVe minimum (0.081) and maximum (0.927) fall within the range between the SALVE minimum (0.05) and maximum (0.984). The mean sharpness of the SALVE dataset is 2.029 Acum, ranging from 1.4 to 3.328 Acum. Similar to the Bioacoustic Index, a higher within STD (0.356 Acum) than between STD (0.305 Acum) can be observed, which is also the case for the Be-MoVe dataset (0.266 & 0.185 Acum resp.).

**Table 1:** Descriptive Statistics for the SALVE and Be-MoVe dataset. Displayed are the arithmetic mean, the minimum (Min.) the maximum (Max.), the standard deviation between the average of each recording location (STD Between) and the average of the standard deviations of the measurements at the respective recording locations (STD Within). The unit of LAeq is dB(A). The unit of sharpness is Acum.

|  | SALVE |  |  |  |  | Be-MoVe |  |  |  |  |
| --- | --- | --- | --- | --- | --- | --- | --- | --- | --- | --- |
|  | Mean | Min. | Max. | STD Between | STD Within | Mean | Min. | Max. | STD Between | STD Within |
| n | 785 | 785 | 785 | 785 | 2,746 | 22 | 22 | 22 | 22 | 90 |
| LAeq | 55.306 | 33.584 | 79.260 | 6.546 | 4.204 | 60.252 | 50.413 | 68.852 | 5.066 | 2.501 |
| Articulation Index (RMS) | 0.846 | 0.207 | 1.000 | 0.136 | 0.076 | 0.700 | 0.447 | 0.942 | 0.146 | 0.051 |
| BIO | 5.882 | 2.425 | 10.433 | 0.985 | 1.190 | 7.424 | 6.454 | 8.694 | 0.824 | 1.249 |
| Link Density | 0.563 | 0.05 | 0.984 | 0.203 | 0.210 | 0.478 | 0.081 | 0.927 | 0.234 | 0.167 |
| Sharpness (Max) | 2.029 | 1.400 | 3.328 | 0.305 | 0.356 | 1.901 | 1.585 | 2.385 | 0.185 | 0.266 |

#### Modelling

##### Feature Reduction

From the 292 initial predictor variables, 215 are identified as unimportant, resulting in a list of 77 LUT categories relevant for predicting acoustic properties in the urban environment (28 LUT categories for 50 m buffer and 49 LUT categories for the 300 m buffer). A list of all relevant LUT categories can be found in the Appendix.

##### Hyperparameter Tuning

We apply a grid search in combination with a 5-fold cross validation to tune the hyperparameters of the gradient boosting model on the test/validation data of the SALVE dataset. We find the optimal parameter for number of trees, maximum tree depth, and learning rate for the indices LAeq (1000, 3, 0.01), Articulation Index (5000, 3, 0.001), Bioacoustic Index (500, 3, 0.01), Link Density (500, 3, 0.01) and Sharpness (5000, 3, 0.001). The results for the performance measures from the final 5-fold cross validation can be found in Table 2. For the LAeq, we find an MAE of 3.88 (STD=0.355) and an RMSE of 4.969 (0.454). The higher RMSE indicates an impact of larger outliers between predictions and measurements. This pattern can be found for each acoustic index, with the RMSE being approximately 1.3 times greater than the MAE. Overall, the R^2^ varies considerably across the models, ranging from a low of 0.133 (0.078) for Sharpness to a high of 0.522 (0.054) for the Articulation Index. The model for the BIO also shows a relatively low explanatory power (0.135 ± 0.104), while for the LAeq, it is located at the upper end (0.463 ± 0.058), and for Link Density, it falls in the mid-range (0.286 ± 0.056). These results indicate that the models show varying predictive quality [38]. In addition, the relatively low STDs in comparison to the means indicate a stability of model performance across different folds.

**Table 2.** Results of the 5-fold cross validation. Displayed are the means ± standard deviations of the respective performance measure; (MAE=Mean Absolute Error, RMSE=Root Mean Square Error, R^2^=Coefficient of Determination). The unit of LAeq is dB(A). The unit of sharpness is Acum.

| | MAE | RMSE | $R^2$ |
| --- | --- | --- | --- |
| LAeq | 3.880 ± 0.355 | 4.969 ± 0.454 | 0.463 ± 0.058 |
| Articulation Index (RMS) | 0.072 ± 0.006 | 0.095 ± 0.006 | 0.522 ± 0.054 |
| BIO | 0.675 ± 0.063 | 0.903 ± 0.065 | 0.135 ± 0.104 |
| Link Density | 0.138 ± 0.006 | 0.171 ± 0.006 | 0.286 ± 0.056 |
| Sharpness (Max) | 0.212 ± 0.015 | 0.277 ± 0.019 | 0.133 ± 0.078 |

#### 3.3. Model Performance

To investigate the model performance on unseen data, we apply the SALVE_Train_ model on the SALVE_Test_ data and the SALVE_All_ model on the Be-MoVe data. Results can be found in Table 3. For a more detailed investigation, we also provide scatter plots and histograms between true values and model predictions as well as SHAP values to investigate the most predictive features (Figure 1).

**Table 3.** Performance measures for model application on test data. For SALVE_Test_ the model trained on the SALVE_Train_ model is used. For Be-MoVE the SALVE_All_ model is used; (MAE=Mean Absolute Error, RMSE=Root Mean Square Error, R^2^=Coefficient of Determination). The unit of LAeq is dB(A). The unit of sharpness is Acum.

|  | SALVE<sub>Test</sub><br>(n = 117) |  |  | Be-MoVE<br>(n = 22) |  |  |
| --- | --- | --- | --- | --- | --- | --- |
|  | MAE | RMSE | R<sup>2</sup> | MAE | RMSE | R<sup>2</sup> |
| LAeq | 3.795 | 4.788 | 0.518 | 3.466 | 4.360 | 0.458 |
| Articulation Index (RMS) | 0.056 | 0.078 | 0.609 | 0.123 | 0.156 | -0.149 |
| BIO | 0.652 | 0.851 | 0.300 | 1.102 | 1.242 | -3.692 |
| Link Density | 0.140 | 0.172 | 0.273 | 0.156 | 0.205 | 0.312 |
| Sharpness (Max) | 0.216 | 0.290 | 0.256 | 0.205 | 0.239 | -0.673 |

**Figure 1.**
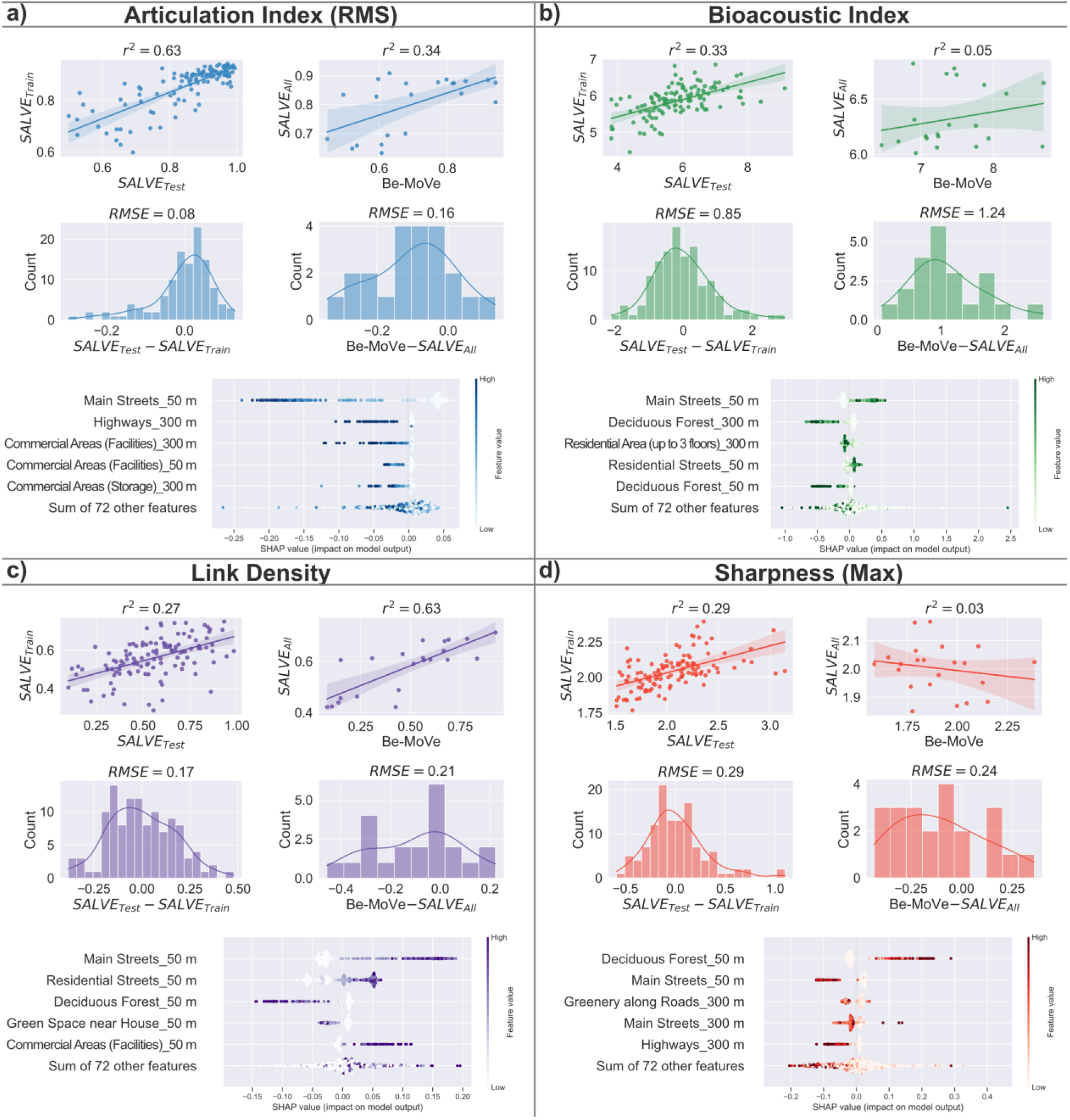
Model performance visualized using scatter plots between true values and model predictions (incl. 95% confidence interval), histograms of the difference between true values and model predictions as well as SHAP values for the top five most important predictor variables in the gradient boosting model for Articulation Index (a), Bioacoustic Index (b), Link Density (c) and Sharpness (d); (RMSE=Root Mean Square Error, r^2^=squared Pearson correlation). The unit of LAeq is dB(A). The unit of sharpness is Acum.

For the LAeq predictions of the SALVE_Test_ dataset, we find slightly lower MAE and RMSE as well as an increased R^2^. This tendency is more pronounced for the model application on the Be-MoVe data, with even lower values for MAE and RMSE. However, all values fall within the results from 5-Fold cross validation (± STD), underlining the robustness of the model for completely unseen LAeq data. For the Articulation Index we find that the model’s performance is comparable to or even better than that of the cross validation when applied to the SALVE_Test_ data, the BIO and the Sharpness. However, its performance decreases strongly when applied to the Be-MoVe data. Here, MAE and RMSE double for the Articulation Index and increase by a factor of approx. 1.5 for the BIO. Although the values slightly decrease for Sharpness, the R^2^ value drops below 0 for all three indices, indicating poor explanatory power in the model’s prediction for the Be-MoVe data. This is underlined by investigating the scatter plots of the linear fit between predicted and measured values (Figure 1). With a r^2^ of < 0.1, the BIO and the Sharpness show close to no substantial linear relation, especially when compared to the model application on the SALVE_Test_ data (r^2^=0.33 and 0.29 resp.). In contrast, the Articulation Index still exhibits a linear but weaker relationship, with the r^2^ decreasing from 0.63 to 0.34. Especially pronounced is the heteroscedastic pattern, which shows a very good fit for higher but an increasingly poor fit for lower Articulation Index values. The performance measures for Link Density remain relatively robust. Results on the SALVE_Test_ data are consistent with those from cross-validation, while the application to the Be-MoVe data shows a slight increase in MAE and RMSE, along with a modest increase in R^2^.

Regarding the predictive importance of LUT categories, SHAP values indicate that “Main Streets” are consistently among the top predictors – ranking as the most important variable for all indices except Sharpness, where it ranks second. Similarly, “Highways” (for Articulation Index and Sharpness) as well as “Residential streets” (for BIO and Link Density) are among the top five most important predictors. Furthermore, different commercial areas play an important role for the AE in regards to Articulation Index and Link Density. The presence of all these LUT almost exclusively increase (BIO & Link Density) or decrease (Articulation Index & Sharpness) the respective indices. The opposite can be observed for the frequently important predictor “Deciduous Forest”. Looking at the distribution by buffer sizes, the immediate surrounding seems to be slightly more important than the broader environment for predicting the acoustic indices, with twelve out of the twenty top five predictors relating to the 50 m Buffers. Here, the Link Density stands out, as all of its most important predictors belong to the 50 m buffer.

#### 3.4. Model Performance in Comparison to the Strategic Noise Map

For an improved understanding of the results and an embedding in the overall context of predicting the environmental AE, we compare the performance of our model in predicting total environmental noise to the performance of the SNM of Bochum (Figure 2). For all investigated measures, we see an overall improved performance of the LUT based models against the SNM (Figure 2a). In direct comparison, the SNM MAE for predicting the LAeq is 1.84 dB(A) higher than that of the SALVE_Train_ model in predicting LAeq measures from SALVE, while the RMSE is 3 dB(A) higher. Considering the R^2^, the SNM estimates show a poor performance in predicting total environmental noise, with a value of -0.307, indicating that the model performs worse than a naive model using the arithmetic mean of the test-data. However, in contrast to the BIO and Sharpness models, the scatter plots (Figure 2b) still reveal a substantial linear relation of r^2^=0.32 between predicted and measured LAeq values. The biggest deviations between measured and predicted LAeq values are found for low predictions around 35 dB(A) and measured values between approx. 40 and 70 dB(A).

**Figure 2.**
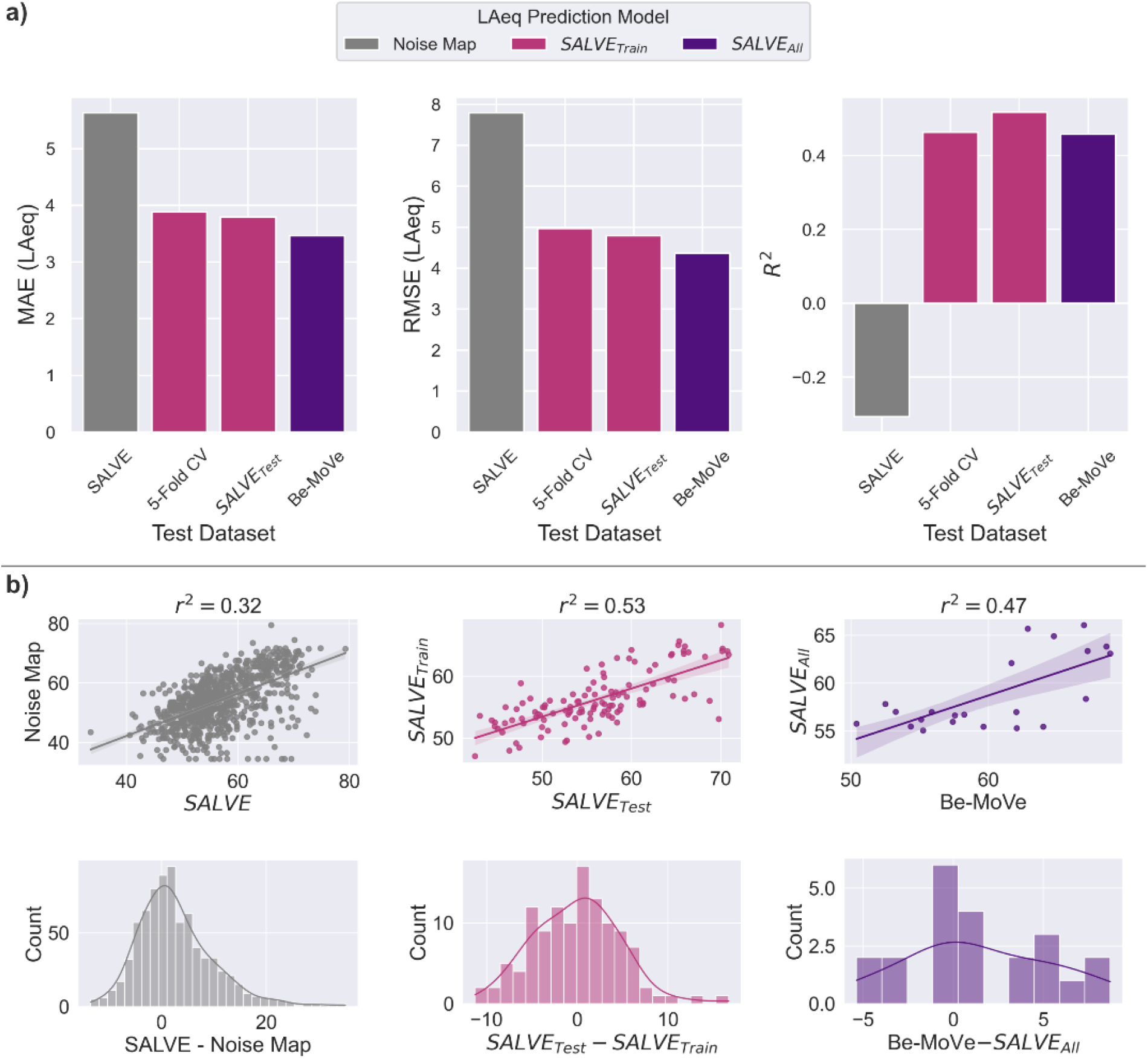
(a) Model performance comparison in predicting total environmental noise, using the strategic noise map, the SALVE_Train_ model and the SALVE_All_ model; (MAE=Mean Absolute Error, RMSE=Root Mean Square Error, R^2^=Coefficient of Determination). (b) Model performance visualized using scatter plots between true values and model predictions (incl. 95% Confidence Interval) as well as histograms of the difference between true values and model predictions; (r^2^=squared Pearson correlation).

This tendency to an underestimation by the predictions is further emphasized by the histogram, which shows a right-skewed distribution of residuals. From there, we see a residual-range from -13.53 dB(A) to 35.07 dB(A). In contrast, residuals are more symmetrically distributed around 0 for the model predictions of the SALVE and Be-MoVe data, ranging from -11.28 to 16.68 dB(A) and -5.37 to 8.72 dB(A). Still, underestimates remain more prevalent also from these models.

## 4. Discussion

The goal of this work is to investigate if LUT based models can predict properties of the urban AE and how they perform in doing so. As LUT data is often widely available, such models offer an efficient approach for estimating AE properties at high spatial resolution – information that is critical for large-scale studies on AE properties beyond noise modelled by SNMs.

For the LUT model applied to predict LAeq, all performance metrics indicate improved performance compared to the predictions from the SNM. We find improved (i.e. decreased) MAE (by approx. 2 dB(A)) and RMSE (3 dB(A)) for the model application on unseen data from two datasets. As an increase of 3 dB(A) corresponds to a doubling in energy, these represent substantial differences between the LUT based model and the SNM. This is underlined by the comparison of R^2^, which even becomes negative for the SNM predictions. However, it should be noted that SNMs are not specifically designed to estimate total environmental noise, but rather noise from specific sound sources (major road, rail and air traffic as well as industry noise). Furthermore, estimates are made for a height of approx. 4 m and at a resolution of 5 × 5 m. In addition, the model exclusion of roads with less than six million vehicle passages a year complicates the use of SNMs for total noise assessment. Although SNM results are often treated as synonymous with total noise pollution, our results should not be viewed as a shortcoming of the SNM. Rather, the results highlight that there are substantial noise sources contributing to total environmental noise, which are not considered by SNMs. This finding is in line with several results from the literature, which also highlight the conceptual shortcoming of SNM when interpreted as total environmental noise [39, 40].

Our findings are largely in line with those of previous studies that modelled comparable noise concepts based on land use data. Liu et al. [14] report a MAE of 3.47 dB(A) and a RMSE of 4.44 dB(A) with an R^2^ of 0.58 in predicting LAeq measurements from five Canadian cities. Aguilera et al. [11] report R^2^ values between 0.66 and 0.87 and also provided comparisons to SNMs, with an r^2^ between 0.38 and 0.61. However, they predict road traffic noise, which represents only a part of the urban AE. In addition, there are several other studies that support our findings [13, 15, 16, 41]. Although these models include additional information to LUTs (e.g. traffic volume, meteorological data) and predict different forms of noise, their performance is close to what we find in our study.

In addition to predict the LAeq, we also predicted the Articulation Index, the BIO, the Link Density and the maximum Sharpness. Here, results are mixed. While for all indices, a moderate to good performance on the SALVE_Test_ data can be observed, performance drops substantially when applied to the Be-MoVe data. This is especially true for the Articulation Index model, which showed the highest R^2^ of all indices (0.609), but became negative for the Be-MoVe data (−0.149). Inspecting the scatter plots, we find that there is still a substantial linear relation between predicted and measured value (r^2^=0.34), though model performance clearly declines at Articulation Index values lower then approx. 0.8. As 68% of the Be-MoVe datasets values are below this value, this might explain the low performance for predictions in more dense and diverse built-up urban environments. Still, the model performs reasonably well in predicting Articulation Index values at locations with a high percentage of intelligible speech. No such patterns nor substantial linear relationships are found for the models predicting the BIO and the Sharpness. In both cases, the R^2^ becomes negative for the Be-MoVe data. For the BIO, the MAE and the RMSE also strongly increase, while they stay approx. the same for the Sharpness. Overall, this indicates that the models incorporating only LUT information are not performing well in predicting these indices. One reason for that could be that the indices focus on frequency power related information, rather than overall sound pressures. Since frequency power tends to vary more over time than across locations [42], a model based on seasonal averages may overlook important temporal dynamics. In addition, the suitability of the BIO and Sharpness indices for capturing meaningful information about the urban AE is still subject to debate [25]. In contrast, the model predicting the Link Density shows similar performance measures between the applications on both datasets. Although the R^2^ values between 0.27 and 0.31 only show a moderate fit, an increased r^2^ from 0.27 to 0.63 for the Be-MoVe dataset indicates its suitability for predicting the AE in more dense and diverse built-up urban environments. While similar concerns regarding frequency information also apply to Link Density, the current results are promising and are likely to improve with models that incorporate temporal information.

To the best of our knowledge, this is the first study to use LUT-based models for predicting acoustic indices that capture acoustic properties beyond noise at a citywide scale, thus, no comparable results are available. However, in comparison to the performance of the SNM, both the models for the LAeq and Link Density provide promising and robust performances for predicting AE properties at a high spatial scale.

### 4.1. Strengths and Limitations

One of the major strengths of this study is the demonstration, that models solely based on readily available LUT information are generally suitable for predicting AE properties. Once the model is built, this enables a fast application for predicting the respective AE properties on high spatial resolution. For example, Figure 3 represents the application of the SALVE_All_ model to predict the LAeq and the Link Density for the research area of three cities (approx. 450 km^2^), which took less than 10 s in computation time for each of the respective indices on a standard office PC (using a 11th Gen Intel(R) Core(TM) i9-11900K at 3.50GHz and 64 GB of RAM). In theory, these models can easily be applied to predict further acoustic properties of interest. Additional strengths are that we compare our results to the performance of the SNM using a comprehensive dataset of acoustic measurements across the city of Bochum from the SALVE project as well as the model evaluation, using two independent test datasets.

**Figure 3.**
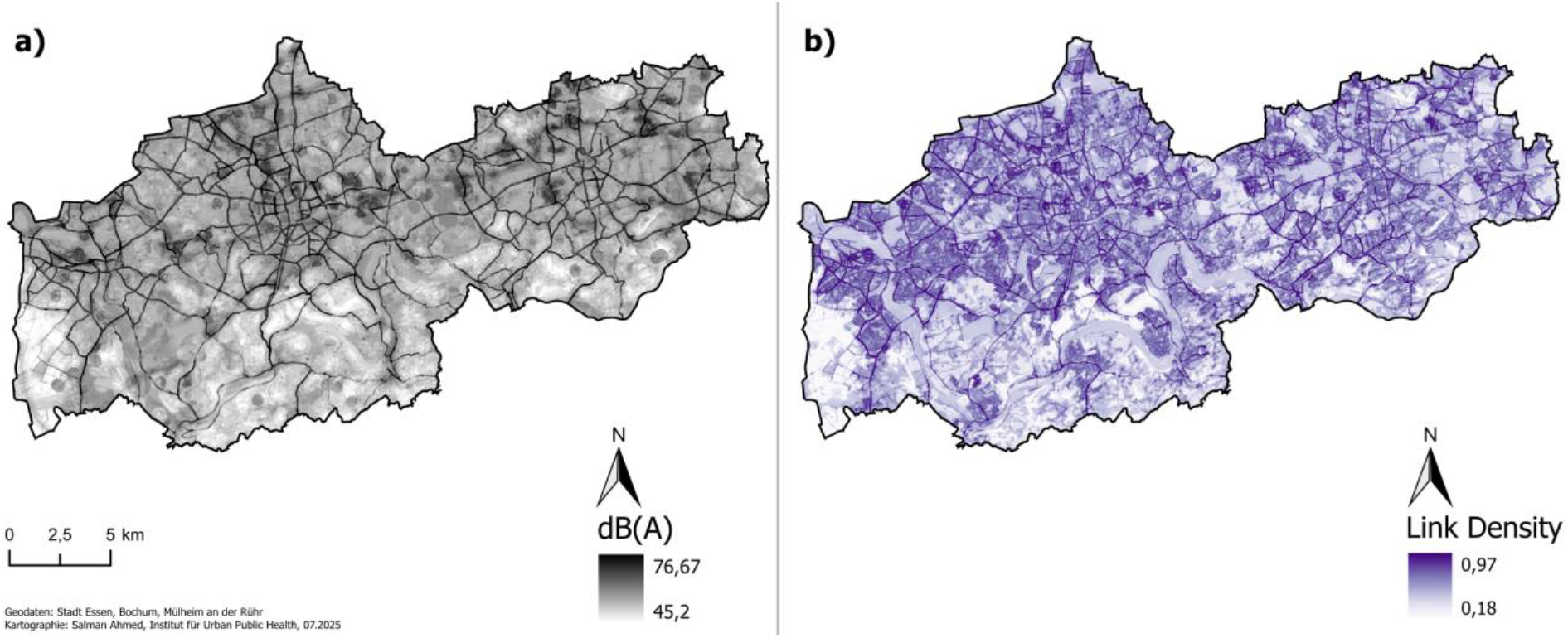
Application of the gradient boosting models to predict total environmental noise (a) and link density (b) for the area of the cities Bochum, Essen and Mülheim an der Ruhr. Calculation time for each map was below 10 s.

However, limitations of this study comprise the already mentioned neglected temporal information, which might be especially important to predict frequency related AE properties. In addition, we only focus on the daytime AE, as measurements are only available between 09:00 and 19:30. As the night-time AE plays an important role for health related issues like sleep [1], our models may not be suitable for accurately estimating exposure levels during this time frame. Another important limitation is that the model is only based on the relations between LUT in the surrounding of acoustic measurements in Bochum, thus, potentially important LUTs (e.g. airports) are not considered in predicting AE properties. Methodologically, we use a gradient boosting model, whose predictions are bound between the minimum and maximum values provided in the training data. While for indices like the Link Density or the Articulation Index, the empirical measures are close to the theoretical bandwidth, the model will fail with predictions for locations with higher or lower values for the other indices. Furthermore, although we performed sensitivity analysis using extreme gradient boosting [43] without observing performance improvements, other models may outperform gradient boosting in the future.

## 5. Outlook

In this work, we demonstrate the predictive power of solely LUT based models to estimate properties of the urban AE at high spatial resolution. As the model for the LAeq outperforms the SNM in predicting total environmental noise and similar performance was found throughout the literature, it represents a promising approach in estimating total environmental noise at high spatial resolution. Although the predictive power for the Link Density model was lower, its results were robust across datasets and it performed best when applied in dense built-up urban environments. Our results are particularly important for epidemiological studies, as the models offer fast large-scale estimates, using readily available information. In the future, these models could be used to estimate exposure to AE properties, which can then be analysed in relation to human health, enabling the investigation of associations already observed in laboratory and field studies within population-based research. Furthermore, future works should investigate whether the inclusion of temporal information along with available environmental data (e.g. satellite imagery) will enhance the predictive power.

## Data Availability

All data produced in the present study are available upon reasonable request to the authors

## Acknowledgements

We would like thank the “Amt für Stadtplanung und Wohnen” of the city of Bochum for providing access to the strategic noise map of Bochum. Their support and cooperation were instrumental in enabling the spatial analyses carried out in this study.

## Funding

This work was supported by the HEAD-Genuit-Foundation [P-21/02-W]. The funding source had no involvement in the study design; in the collection, analysis and interpretation of data; in the writing of the report; and in the decision to submit the article for publication.

## Notes

### Competing Interest Statement

The authors have declared no competing interest.

